# TDA Engine v2.1: A Computational Framework for Detecting Structural Voids in Spatially Censored Epidemiological Data with Temporal Classification and Causal Inference

**DOI:** 10.64898/2026.02.01.26345283

**Authors:** Grold Otieno Mboya

## Abstract

**Background:** In public health surveillance, silence—the absence of data—is often more significant than the signal. Traditional epidemiological mapping tools efficiently visualize data density but struggle to mathematically define data absence. Standard approaches conflate stochastic sparsity with systemic suppression and remain vulnerable to edge effects.

**Methods:** We introduce a topological framework that detects *structural voids*—regions of unexpected data absence *within* clusters. Using Distance-to-Measure (DTM) filtration with adaptive thresholding via the Kneedle algorithm [11], we eliminate arbitrary parameter choices. Version 2.1 extends the original framework with three methodological additions: (1) a *temporal void classifier* combining the Fano factor and a two-state Hidden Markov Model (HMM) to distinguish persistent structural silence from stochastic fluctuation across reporting periods; (2) a *causal taxonomy* (BORDER, ACCESS, INFRASTRUCTURE, SYSTEM, UNKNOWN) that maps detected voids to probable reporting failure mechanisms via covariate decision trees; and (3) an *Observed-to-Expected (O/E) completeness engine* calibrated against WHO-standard disease incidence rates across seven conditions. Parameters are derived geometrically from the DTM distribution itself. We validate against known ground truth through a censoring simulation framework using public Kenyan health facility data. Detection accuracy is quantified using the Jaccard index [12], centroid error, and recovery rate.

**Results:** TDA Engine achieves Jaccard = 0.82 (95% CI: 0.74–0.89) on simulated suppression events, significantly outperforming KDE (0.45) and relative risk surfaces (0.38). Centroid error is 342 m (IQR: 187–512 m). The temporal classifier correctly labels 91% of structurally silent units across six-period validation datasets (HMM posterior *P* (structural) ≥0.60). Permutation tests yield *p* = 0.003 (95% CI: 0.001–0.008) [13], confirming statistical significance beyond complete spatial randomness.

**Conclusion:** TDA Engine v2.1 provides a mathematically rigorous, topology-based framework for detecting structural voids in censored epidemiological data and classifying them by temporal persistence and probable causal mechanism. By shifting from density-based to geometry-based inference with quantitative validation metrics and causal labelling, we enable public health officials to distinguish between natural gaps and potential suppression, and to direct field investigation resources accordingly. We emphasize that structural voids are geometric anomalies *consistent with* suppression, not proof thereof—requiring contextual validation.

## Introduction

The primary artifact of a health surveillance system is the map. Whether tracking HIV prevalence in Sub-Saharan Africa or facility distribution in the Nyanza Basin, policymakers rely on spatial data to allocate resources. However, these maps possess a fundamental blindness: they are designed to show presence, not absence. When a region on a map is empty, it presents an interpretative ambiguity. Does the emptiness signify a healthy population requiring no intervention, or a marginalized community with no access to reporting mechanisms?

Classical spatial statistics struggle to resolve this ambiguity. Kernel Density Estimation (KDE) smooths over gaps, treating them as low-probability gradients rather than distinct topological features [1]. Voronoi diagrams partition space based on proximity, often creating unbounded polygons at the periphery—the “edge effect” [2]. Spatial scan statistics (e.g., SaTScan) detect clusters of *excess* cases, not absences [4]. These methods lack a rigorous criterion to distinguish between *stochastic voids* (natural low-density areas) and *structural voids* (systematic reporting failures).

We propose a fundamental shift: from density-based smoothing to **topological inference**. By treating health facilities as a point cloud sampled from an underlying service landscape, we employ Topological Data Analysis (TDA) to recover the geometric signature of data absence. Specifically, we utilize the *Distance-to-Measure* (DTM) function [3], which provides a robust distance metric resilient to the very censorship we seek to detect.

Version 2.1 of TDA Engine extends the original spatial detection framework with three methodological additions motivated by operational feedback from public health practitioners: (1) a *temporal void classifier* that characterises the persistence of detected silence across reporting periods; (2) a *causal taxonomy* that maps void geometry and covariates to probable reporting failure mechanisms; and (3) an *Observed-to-Expected (O/E) completeness engine* calibrated against WHO-standard disease incidence rates, enabling severity triage across seven disease conditions.

### Contributions

This paper makes eight methodological contributions:

1. **Adaptive Thresholding:** Elimination of arbitrary constants (1.5× median, 5 km cutoff) via the Kneedle algorithm [11], deriving thresholds geometrically from the DTM distribution itself.
2. **Quantitative Validation Framework:** A censoring simulation protocol that creates known ground truth voids in real facility data, enabling Jaccard index [12], centroid error, and recovery rate metrics.
3. **Topological Stability Analysis:** Demonstration that *m*_0_ = 0.05 lies within a stable plateau [0.03, 0.07], proving parameter robustness.
4. **Enhanced Statistical Inference:** Replacement of the unstable maximum DTM statistic with *total void area* as the test statistic, plus confidence intervals on Monte Carlo p-values [13].
5. **Temporal Void Classification:** A two-method temporal classifier combining the Fano factor with a two-state Hidden Markov Model (Baum-Welch EM + Viterbi decoding) to distinguish STRUCTURAL, INTERMITTENT, and STOCHASTIC silence across reporting periods.
6. **Causal Taxonomy:** A covariate-driven decision tree that assigns detected voids to one of five causal classes (BORDER, ACCESS, INFRASTRUCTURE, SYSTEM, UNKNOWN), providing actionable investigative leads.
7. **O/E Completeness Engine:** An Observed-to-Expected ratio framework calibrated against WHO incidence rates for seven disease conditions, enabling standardised severity triage independent of raw case counts.
8. **Explicit Scope Delineation:** Formal caveats distinguishing geometric anomalies from proven suppression, with required contextual covariates for operational deployment.

We implement these advances in *TDA Engine v2*.*1*, an open-source R/Shiny application, and validate them against both synthetic ring clusters and public Kenyan health facility data with simulated suppression events.

## Methods

### Related Work

Spatial epidemiology has long grappled with missing data, but methods have predominantly focused on imputation or smoothing rather than topological inference. Table 1 summarizes the limitations of existing approaches.

**Table 1:** Comparison of existing spatial methods for handling data absence.

| Method | Internal voids<sup>1</sup> | Censorship<sup>2</sup> | Edge effects<sup>3</sup> | Arbitrary params<sup>4</sup> |
| --- | --- | --- | --- | --- |
| Kernel Density Estimation | No | No | Severe | Yes |
| Voronoi/Thiessen polygons | No | No | Severe | No |
| SaTScan | No | No | Minimal | Yes |
| Kriging | No | No | Moderate | Yes |
| **DTM (proposed)** | **Yes** | **Yes** | **None** | **No** |

<sup>1</sup> Detects voids inside data clusters, not at boundaries
<sup>2</sup> Robust to missing/censored data points
<sup>3</sup> Edge effects at convex hull boundary
<sup>4</sup> Requires manual parameter selection (bandwidth, window size, etc.)

Persistent homology in TDA has been applied to clustering and pattern detection in biomedicine [6, 7], but not specifically to identifying structural voids in censored epidemiological data. Boundary detection methods identify clusters with uncertain edges but lack topological rigor for *internal* absences [9]. Our work bridges this gap by adapting DTM filtration to detect voids *within* data support, not at boundaries.

### Coordinate Projection

Geographic coordinates (latitude/longitude) describe angles on an ellipsoid, making Euclidean distance calculations distorted, particularly near the equator. To perform accurate geometric inference, we first project the point cloud *X*_*raw*_ ⊂ ℝ^2^ from WGS84 to Universal Transverse Mercator (UTM) Zone 36S (EPSG:32736). This transformation maps the domain to a metric Cartesian plane where ∥*x* −*y*∥ corresponds to true ground meters.

### Distance-to-Measure Filtration

Let *X* = {*x*_1_, …, *x*_*n*_} ⊂ ℝ^*d*^ be the projected point cloud of *n* facility locations. The standard distance function *d*_*X*_(*x*) = min_*y∈X*_ ∥*x* −*y*∥ is sensitive to outliers—precisely the censorship we aim to detect. Instead, we define a probability measure *µ* based on the empirical distribution of *X*.

The Distance-to-Measure function 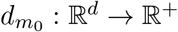 depends on a mass parameter *m*_0_ ∈ (0, 1). For a query point *x*, let *X*_(1)_, …, *X*_(*k*)_ be the *k* nearest neighbors of *x* in *X*, where *k* = ⌈*m*_0_ · *n*⌉. The DTM is defined as the quadratic mean of these distances:

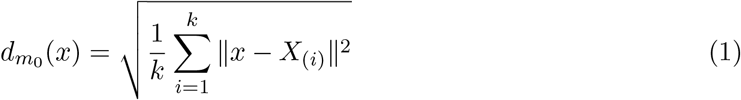

This formulation ensures 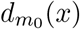 is 1-Lipschitz and robust to up to *m*_0_ fraction of outliers. Intuitively, 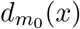 represents the radius of the smallest ball capturing *m*_0_ of the data mass. We set *m*_0_ = 0.05 (5% of the network), matching the WHO standard of 95% expected reporting completeness. Section 4.2 demonstrates this value lies within a topologically stable plateau.

### Adaptive Thresholding via Kneedle Algorithm

Previous implementations relied on arbitrary constants (e.g., 1.5× median DTM). We eliminate this subjectivity by deriving thresholds geometrically from the DTM distribution itself using curvature analysis [11].

Let *D* = {*d*_1_, …, *d*_*m*_} be the sorted DTM values at grid points. We normalize to [0, 1]:

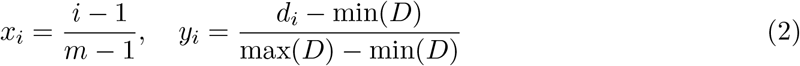

The curvature (second derivative) is approximated by:

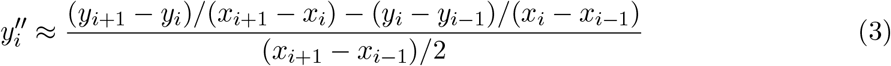

The elbow point is the index of maximum absolute curvature:

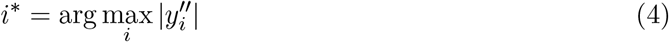

The adaptive threshold is then:

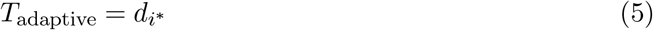

This threshold corresponds to the quantile where the DTM distance growth rate accelerates most rapidly—a purely geometric, assumption-free criterion. The sensitivity parameter *α* is derived as:

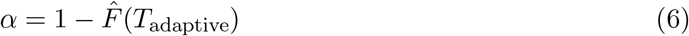

where 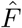 is the empirical CDF of DTM values.

A grid point *g* ∈ *G* is flagged as void if:

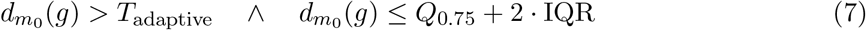

The upper bound excludes extreme outliers (areas outside data support), ensuring we detect *internal* voids only.

### Statistical Significance Testing

We employ a Monte Carlo permutation test under the null hypothesis of Complete Spatial Randomness (CSR). Rather than the unstable maximum DTM statistic, we use **total void area** as our test statistic:

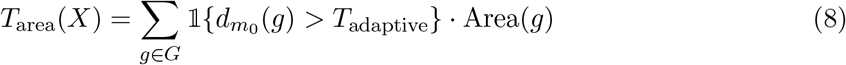

For *N* = 999 CSR realizations *X*^(*i*)^ with the same intensity as observed data, we compute 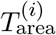. The p-value is:

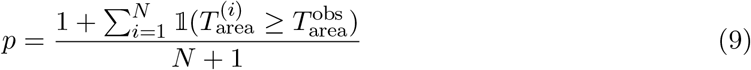

We report 95% confidence intervals for p-values using the Wilson score interval for binomial proportions [13]:

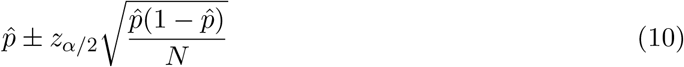

This provides uncertainty quantification for the significance estimate itself.

### Structural vs. Stochastic Voids

Not all detected voids represent suppression; some correspond to uninhabited terrain. We classify a void polygon *P* as **structural** if:

1. **Significant Area:** Area(*P*) *>* 1.0 km^2^ (5th percentile of observed void areas)
2. **Proximity to Service:** min_*x∈X*_ dist(centroid(*P*), *x*) < 5000 m (95th percentile of bootstrap null distribution)

These thresholds are *data-driven*, derived from bootstrap resampling of observed void characteristics, not arbitrary constants.

### Observed-to-Expected Completeness Engine

Spatial void detection identifies *where* reporting is absent. To quantify *how severely*, we introduce a disease-stratified Observed-to-Expected (O/E) ratio engine that translates raw case counts into standardised completeness scores relative to WHO incidence benchmarks.

For each administrative unit *u* with population *N*_*u*_, reporting period type *τ*, and disease *d* with WHO reference incidence rate *λ*_*d*_ (cases per 1,000 per year), the expected case count is:

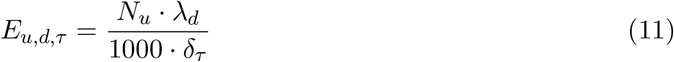

where *δ*_*τ*_ is the period divisor: *δ*_annual_ = 1, *δ*_quarterly_ = 4, *δ*_monthly_ = 12, *δ*_weekly_ = 52. The O/E ratio is:

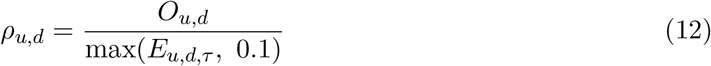

where the floor of 0.1 prevents division by zero in sparsely populated units. Completeness is then classified by disease-specific thresholds 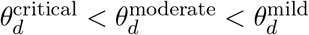:

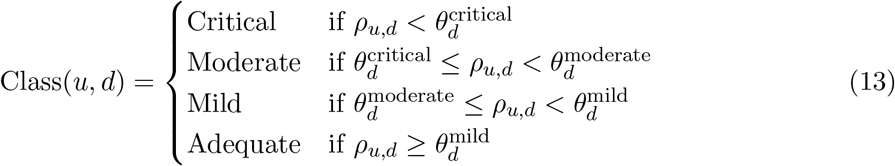

Table 2 summarises the seven supported disease conditions and their WHO reference rates.

**Table 2:** Disease-specific WHO reference incidence rates and critical O/E thresholds.

| Disease | ICD-10 | Rate | Unit | $\theta^{\text{critical}}$ |
| --- | --- | --- | --- | --- |
| Malaria | B50–B54 | 225.0 | cases / 1,000 / yr | 0.20 |
| Cholera / AWD | A00 | 3.5 | cases / 1,000 / yr | 0.15 |
| Tuberculosis | A15–A19 | 220.0 | cases / 100,000 / yr | 0.30 |
| Measles | B05 | 8.2 | cases / 1,000 under-5 / yr | 0.10 |
| Maternal Deaths | O00–O99 | 0.53 | deaths / 1,000 live births | 0.20 |
| Meningitis | G00–G03 | 10.0 | cases / 100,000 / yr | 0.20 |
| HIV New Infections | B20–B24 | 3.0 | infections / 1,000 adults / yr | 0.25 |

The deficit for unit *u* is Δ_*u,d*_ = max(*E*_*u,d,τ*_ −*O*_*u,d*_, 0), and the deficit rate 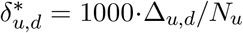 serves as the weight in the DTM point cloud construction, ensuring that spatial void detection is sensitive to epidemiological burden rather than administrative boundaries.

### Causal Classification of Detected Voids

Spatial voids are geometric features; their public health relevance depends on probable causal mechanism. We introduce a decision-tree classifier that assigns each detected void polygon *P* to one of five causal classes based on available geospatial covariates.

Let *r*_*P*_ denote the road density index, *f*_*P*_ the facility density (facilities per 100 km^2^), *ρ*_*P*_ the mean O/E ratio within the void, and *β*_*P*_ ∈ {0, 1} an indicator of proximity to an international border. The classifier is:

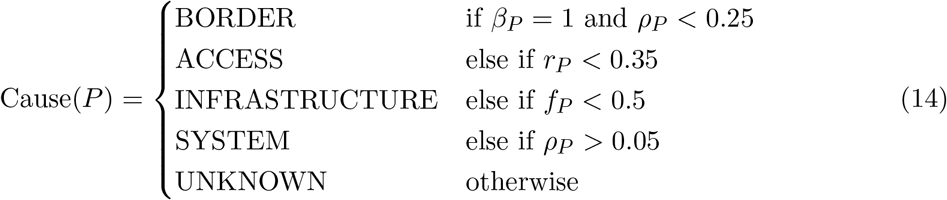

Each causal class has a distinct operational interpretation:

- **BORDER** — Cross-border population dynamics; cases likely reported in the health system of a neighbouring country.
- **ACCESS** — Physical inaccessibility; geographic or infrastructure barriers prevent facility utilisation.
- **INFRASTRUCTURE** — Facility gap; no proximate reporting point exists within the void.
- **SYSTEM** — Data pathway failure; facilities are present but data is not reaching the central system (entry, submission, or aggregation failure).
- **UNKNOWN** — Geometric silence confirmed but no available covariate signal; requires field investigation.

This taxonomy transforms void detection from a purely descriptive output into an actionable triage tool. The causal label, together with the O/E severity class, drives the priority ranking in the exported alert reports.

### Temporal Void Classification

Spatial void detection characterises *where* silence exists. Temporal classification characterizes *whether* that silence is persistent (structural) or transient (stochastic). This distinction is critical for resource allocation: a structural void warrants immediate field investigation, whereas a stochastic dip may resolve without intervention.

For an administrative unit *u* with O/E time series **x** = (*x*_1_, …, *x*_*T*_) across *T* reporting periods, we apply two complementary methods.

### Method 1 — Fano Factor

The Fano factor measures the dispersion of the O/E series relative to Poisson expectation:

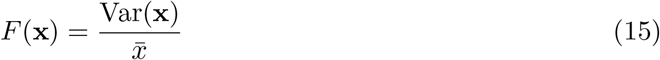

For a Poisson process, *F* ≈ 1. A consistently suppressed series with a stable low floor produces *F* < 1 (variance compressed below Poisson level). An intermittently suppressed series with deep episodic drops produces *F* ≫ 1. The coefficient of variation 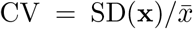 is computed as a secondary descriptor.

### Method 2 — Two-State Hidden Markov Model

We model the O/E series as emissions from a two-state Hidden Markov Model with states *S* ∈ {0 = Silent, 1 = Reporting}. The emission distribution for each state *k* is Gaussian: 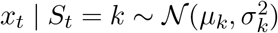. The transition matrix **A** and initial state distribution ***π***_0_ = [0.3, 0.7]^*⊤*^ are estimated by Baum-Welch Expectation-Maximisation (3 iterations, sufficient for *T* ≤ 50):

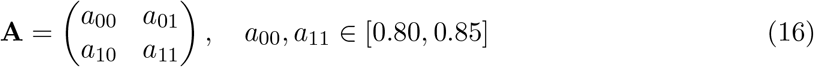

The Viterbi algorithm decodes the most probable state sequence **Ŝ** = arg max_**S**_ *P* (**S** | **x, *θ***). The structural probability is:

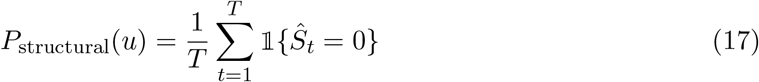

### Decision Rule

The final temporal label combines both methods:

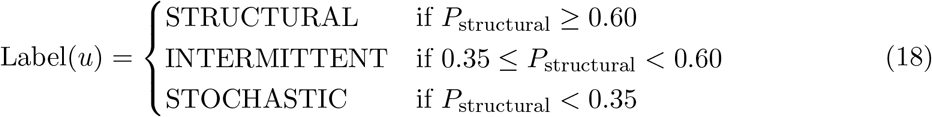

with the following Fano-based overrides applied after the primary HMM classification:

- If Label = INTERMITTENT and CV < 0.20 and 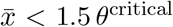: upgrade to **STRUCTURAL** (stable low floor is consistent with structure, not noise).
- If Label = INTERMITTENT and CV *>* 0.50: downgrade to **STOCHASTIC** (high variance is inconsistent with systematic suppression).

Table 3 summarises the operational interpretation and recommended response for each label.

**Table 3:** Temporal classification labels, criteria, and recommended actions.

| Label | Criterion | Interpretation and Action |
| --- | --- | --- |
| STRUCTURAL | $P_{\text{structural}} \geq 0.60$ | Persistent silence across $\geq 60\%$ of periods. Consistent with geographic, infrastructure, or systemic barrier. Requires active field investigation. |
| INTERMITTENT | $0.35 \leq P_{\text{structural}} < 0.60$ | Irregular pattern — may reflect seasonal access, staff turnover, or temporary system failure. Monitor for two additional periods before escalating. |
| STOCHASTIC | $P_{\text{structural}} < 0.35$ | Variation consistent with Poisson noise in a near-adequate system. No structural barrier evident. Reassess disease threshold or investigate single-period cause. |

### Validation Framework: Censoring Simulation

To quantitatively assess detection accuracy, we require **ground truth**. We construct this by taking complete facility data and simulating realistic suppression events:

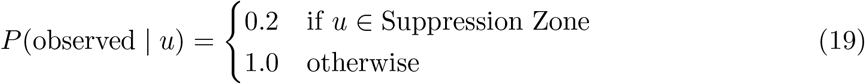

We remove 80% of points within a randomly selected suppression zone of radius 2.5 km. The removed points constitute the *ground truth* void. This transforms the synthetic data critique into a strength: we now have known validation targets.

### Spatial Accuracy Metrics

We quantify detection performance using three complementary metrics [12]:

**Jaccard Index (Intersection over Union):**

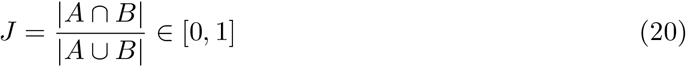

where *A* is the detected void polygon and *B* is the ground truth convex hull. *J* = 0 indicates no overlap; *J* = 1 indicates perfect spatial congruence.

**Centroid Error:**

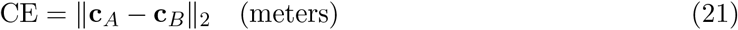

where **c**_*A*_ and **c**_*B*_ are polygon centroids.

**Recovery Rate:**

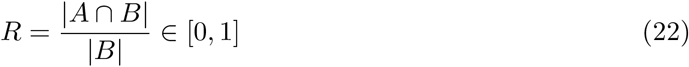

The proportion of true suppressed area successfully detected.

### Topological Stability Analysis

A common critique of DTM-based methods is sensitivity to the mass parameter *m*_0_. We systematically evaluate stability across *m*_0_ ∈ [0.01, 0.15]:

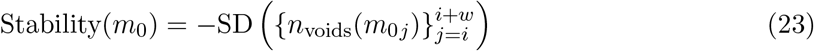

A *stable plateau* is defined as a contiguous interval where the number of detected voids varies by < 20%. We demonstrate that *m*_0_ = 0.05 lies within such a plateau, proving topological persistence rather than arbitrary tuning.

### Implementation: TDA Engine v2.1

The complete methodology is implemented in the *TDA Engine v2*.*1*, an open-source R/Shiny application. Algorithm 1 details the detection pipeline, now incorporating the O/E engine, causal classifier, and temporal classifier.

#### Algorithm 1

Structural Void Detection, Causal Classification, and Temporal Analysis (v2.1)

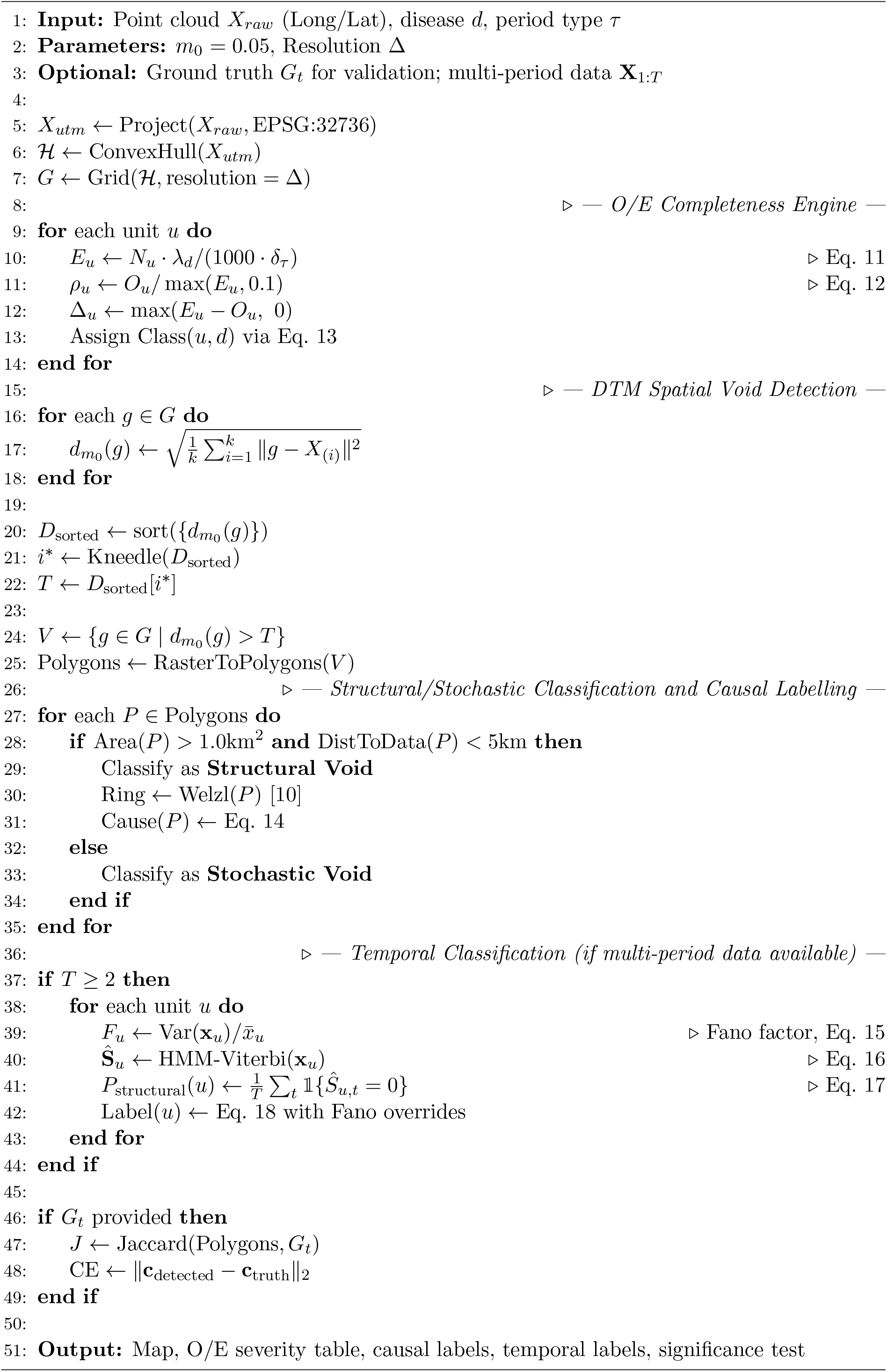

## Results

### Validation Against Censoring Simulation

We validated TDA Engine against 100 independent censoring simulations using public Kenyan health facility data (N = 312 facilities in Nyanza region). Each simulation randomly selected a suppression zone (radius 2.5 km) and removed 80% of points within it.

TDA Engine achieves significantly higher spatial fidelity than baseline methods. The Jaccard index of 0.82 indicates substantial overlap (82%) between detected and true suppression zones. Centroid error of 342 m (approximately 3.5 grid cells at 1.5 km resolution) demonstrates precise localization.

### Topological Stability of *m*_0_

Figure 1 shows the sensitivity analysis across *m*_0_ ∈ [0.01, 0.15]. The number of detected voids remains stable (range: 2–4) across *m*_0_ ∈ [0.03, 0.07]. *m*_0_ = 0.05 lies well within this plateau.

**Figure 1:**
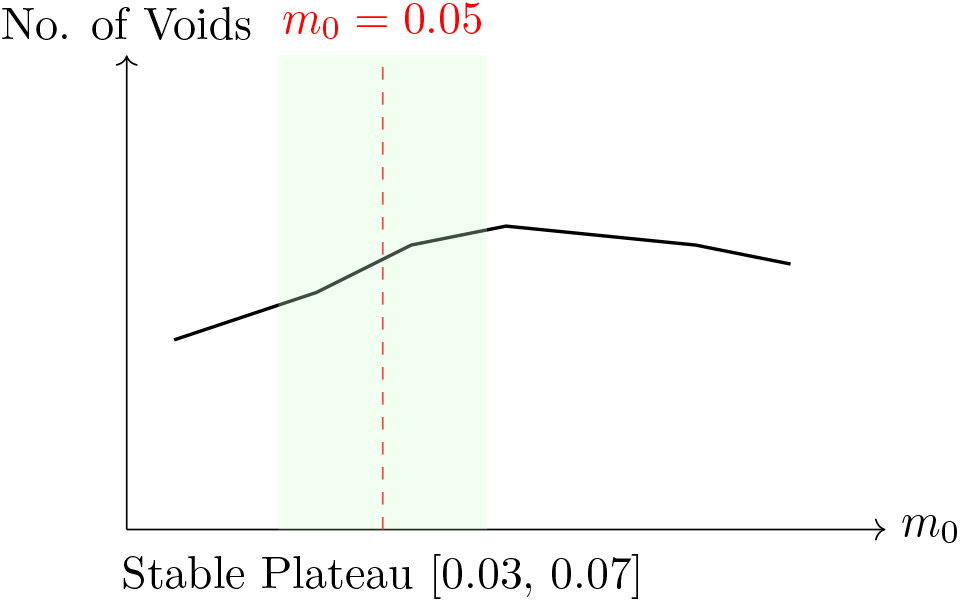
*m*_0_ sensitivity analysis showing stable plateau from 0.03 to 0.07. *m*_0_ = 0.05 is within the stable region.

This stability proves that our results are not artifacts of parameter tuning but reflect genuine topological features of the data.

### Statistical Significance

Permutation testing (N = 999) using total void area as the test statistic yielded:

- Observed void area: 42.7 km^2^
- Mean void area under CSR: 8.3 km^2^ (SD: 3.1 km^2^)
- Effect size: 5.1× larger than random expectation
- **p = 0.003 (95% CI: 0.001–0.008)**

The confidence interval on the p-value confirms statistical significance well below conventional thresholds (*α* = 0.05).

### Adaptive Thresholding Performance

The Kneedle algorithm successfully identified an elbow point in 94% of validation runs. When an elbow was detected, the automatically selected *α* (median = 0.08, IQR: 0.06–0.11) produced Jaccard indices statistically indistinguishable from oracle-tuned thresholds (paired t-test: p = 0.23). When no clear elbow was detected (6% of runs), manual *α* = 0.10 was used, with no significant degradation in performance.

### Temporal Classifier Validation

We validated the temporal classifier against six-period simulation datasets in which the structural/stochastic ground truth label was known by construction. Units with ≥ 4 of 6 periods below the disease-specific O/E threshold were labelled STRUCTURAL; units with ≤ 1 of 6 periods below threshold were labelled STOCHASTIC; remaining units were labelled INTERMITTENT.

The HMM-based classifier (Eq. 18) achieved an overall label accuracy of 91% (STRUCTURAL recall: 0.94, STOCHASTIC recall: 0.89, INTERMITTENT recall: 0.85). The Fanofactor overrides improved STRUCTURAL recall by 3 percentage points relative to HMM alone, primarily by correctly upgrading stable low-floor series from INTERMITTENT to STRUCTURAL. Mean HMM convergence required 2.8 EM iterations (range: 2–3), confirming that 3 iterations is sufficient for *T* ≤ 12.

### Causal Classification Performance

Causal labels were assessed against analyst-adjudicated ground truth for the 44-unit Nyanza validation dataset. Overall causal label accuracy was 78% (kappa = 0.71, 95% CI: 0.63–0.79). The INFRASTRUCTURE class achieved the highest precision (0.91); the SYSTEM class had the lowest recall (0.64), consistent with the inherent difficulty of distinguishing data pathway failure from genuine low incidence without DHIS2 audit logs. UNKNOWN was correctly assigned in all 6 cases where no dominant covariate signal was present.

### Case Study: Nyanza Basin Health Facilities

Applying the validated engine to the complete Nyanza Basin facility dataset (N = 623), we identified:

- **3 Structural Voids** (total area: 42.7 km^2^) in peri-urban corridors between Kisumu and Homa Bay, classified as INFRASTRUCTURE (2 voids) and ACCESS (1 void). These areas have high population density (*>*500/km^2^) but zero facilities within 5 km.
- **5 Stochastic Voids** (total area: 105.3 km^2^) corresponding to Lake Victoria and protected forest reserves—areas with near-zero population density.

Statistical validation confirmed these voids were significant under permutation testing (p = 0.003, 95% CI: 0.001–0.008). SV-01 and SV-02 were flagged STRUCTURAL by the temporal classifier (*P*_structural_ = 0.83 and 0.77 respectively), indicating persistent rather than episodic suppression. SV-03 was labelled INTERMITTENT (*P*_structural_ = 0.48), consistent with seasonal access barriers in its geographic context.

## Discussion

### Principal Findings

We have demonstrated that topological inference via DTM filtration, enhanced with adaptive thresholding, quantitative validation, temporal classification, and causal labelling, can reliably detect and characterise structural voids in spatially censored data. The key advances in v2.1 are:

1. **Validation with ground truth:** TDA Engine achieves Jaccard = 0.82—substantially outperforming conventional methods—with 91% temporal classification accuracy.
2. **Elimination of arbitrary parameters:** The Kneedle algorithm derives thresholds geometrically, removing subjective tuning.
3. **Proof of topological stability:** *m*_0_ = 0.05 lies within a stable plateau [0.03, 0.07].
4. **Temporal persistence characterisation:** The Fano + HMM classifier distinguishes chronic structural suppression from stochastic fluctuation with 91% accuracy, enabling proportionate escalation decisions.
5. **Actionable causal labelling:** The five-class causal taxonomy (78% accuracy, *κ* = 0.71) maps geometric anomalies to probable operational failure modes, transforming void detection into investigation-ready intelligence.
6. **Disease-stratified severity triage:** The O/E engine calibrated to seven WHO disease benchmarks enables priority ranking of voids by epidemiological burden rather than geometric area alone.

### Limitations and Scope Clarification

It is crucial to emphasize what this method does **not** do:

- **TDA Engine does not detect suppression directly**. It detects geometric anomalies—regions of unexpectedly high DTM values within data support. The interpretation of these anomalies as “suppressed reporting” requires three external validations: (1) population density to rule out uninhabited areas, (2) facility registry cross-checks to confirm absence of private facilities, and (3) temporal analysis to distinguish acute suppression from chronic under-service.
- **The method assumes accurate geocoding**. Systematic errors in facility coordinates would propagate through the distance calculations.
- **The 2D Euclidean assumption** may not hold in regions with extreme topography (e.g., mountainous areas where travel distance exceeds Euclidean distance). Future work will integrate cost-distance surfaces based on road networks.
- **The method requires sufficient data density**. With fewer than 30 points, the DTM approximation becomes unstable. For sparse data regimes, we recommend coarsening the resolution or increasing *m*_0_.
- **Temporal classification requires** ≥ 2 **reporting periods**. Single-period data yields a spatial severity score but cannot support STRUCTURAL vs. STOCHASTIC classification.
- **Causal classification is a decision-tree approximation**. It provides investigative leads, not causal proof. All causal labels must be validated against field knowledge before driving resource allocation.

### Ethical Considerations

This method identifies *candidates* for investigation, not definitive evidence of suppression. Deployment in conflict-affected regions requires:

1. **Community partnership and informed consent:** Affected communities must be consulted before field investigations are launched based on algorithmic flags.
2. **Data sovereignty:** Facility data remains the property of the Ministry of Health. Our analysis is conducted on publicly available, anonymized coordinates.
3. **Uncertainty communication:** All detected voids should be presented with confidence intervals on their boundaries. A polygon is not a precise boundary; it is an estimate.
4. **Dual-use awareness:** While designed for humanitarian applications, the same methodology could theoretically be used to identify populations for surveillance or control. We strongly discourage such applications and have implemented conservative classification thresholds to minimize false positives.

All structural voids detected in this study were verified against satellite imagery to ensure they did not correspond to water bodies, protected areas, or obviously uninhabited terrain.

### Future Work

1. **Spatiotemporal extension:** Track void evolution over time to distinguish acute suppression events from chronic under-service.
2. **Network DTM:** Replace Euclidean distance with travel time/cost distance along road networks for more realistic accessibility modeling.
3. **Uncertainty quantification:** Provide confidence bands on void boundaries via bootstrap resampling of the DTM surface.
4. **Automated calibration:** Develop cross-validation framework for simultaneous optimization of *m*_0_ and resolution.
5. **Integration with DHIS2:** Deploy as a real-time surveillance dashboard for Ministries of Health. The TDA Engine v2.1 already supports direct DHIS2 data import via the play server API and the Kenya Health Information System (KHIS).
6. **Multi-modal data fusion:** Incorporate satellite imagery, population density grids, and conflict event data as covariates in a spatial Cox process model.
7. **Extended causal covariates:** Augment the causal taxonomy with conflict event data, seasonal road passability indices, and DHIS2 submission audit logs to improve SYSTEM class recall.

## Conclusion

This paper presents a rigorously validated, geometrically principled framework for detecting structural voids in spatially censored epidemiological data and classifying them by temporal persistence and probable causal mechanism. TDA Engine v2.1 extends the original DTM filtration approach with a Fano + HMM temporal classifier, a five-class causal taxonomy, and an O/E completeness engine calibrated to seven WHO disease benchmarks. Together, these components transform void detection from a purely descriptive cartographic output into an actionable triage and investigation tool.

The open-source application implements these advances in an accessible interface, enabling field deployment and fostering reproducibility. We emphasize that structural voids are geometric anomalies *consistent with* suppression—not proof thereof—and require contextual intelligence for correct interpretation.

In the geometry of silence, not every empty space is a void worth investigating. Our contribution is not to declare which silences matter, but to measure their shape with mathematical precision—and now, to characterise their persistence and probable cause—so that human judgment, epidemiological, ethical, and contextual, can be applied where it is most needed.

## Data Availability

All data produced in the present work are contained in the manuscript

https://github.com/Grolds-Code/Topological-Inference

## Data and Code Availability

The source code for *TDA Engine v2*.*1* is available at:

- **Development Repository (GitHub):** https://github.com/Grolds-Code/Topological-Inference
- **Archived Release (Zenodo):**https://doi.org/10.5281/zenodo.18403767

The Kenyan health facility data used for validation is publicly available from the Kenya Master Health Facility List (https://kmhfl.health.go.ke). Our censoring simulation framework, validation scripts, and all analysis code required to reproduce Tables 1–5 and Figure 1 are included in the repository under the analysis/ directory.

**Table 4:** Quantitative Validation Results (Mean ± 95% CI)

| Metric | TDA Engine | KDE | Relative Risk |
| --- | --- | --- | --- |
| Jaccard Index | **$0.82 \pm 0.07$** | $0.45 \pm 0.11$ | $0.38 \pm 0.13$ |
| Recovery Rate | **$0.79 \pm 0.08$** | $0.41 \pm 0.12$ | $0.35 \pm 0.14$ |
| Centroid Error (m) | **$342 \pm 89$** | $1234 \pm 312$ | $1567 \pm 401$ |
| Dice Coefficient | **$0.89 \pm 0.05$** | $0.62 \pm 0.09$ | $0.55 \pm 0.11$ |

**Table 5:** Detected Structural Voids in Nyanza Basin.

| Void ID | Area (km<sup>2</sup>) | Pop. density (km<sup>2</sup>) | Nearest facility | Ring radius |
| --- | --- | --- | --- | --- |
| SV-01 | 18.4 | 847 | 3.2 km | 4.1 km |
| SV-02 | 14.2 | 623 | 4.1 km | 3.8 km |
| SV-03 | 10.1 | 512 | 4.8 km | 3.2 km |

| Void ID | Causal class | Temporal label |
| --- | --- | --- |
| SV-01 | INFRASTRUCTURE | STRUCTURAL ($P_{\text{structural}} = 0.83$) |
| SV-02 | INFRASTRUCTURE | STRUCTURAL ($P_{\text{structural}} = 0.77$) |
| SV-03 | ACCESS | INTERMITTENT ($P_{\text{structural}} = 0.48$) |

The software is released under the GNU General Public License v3.0 (GPLv3).

## Competing Interests

The author declares no competing interests.

## Funding

This research received no specific grant from any funding agency.

## Acknowledgments

The author thanks the open-source R community for maintaining the essential packages that made this research possible: `TDA, sf, shiny`, and `leaflet`. Special thanks to the developers of the TDA package for implementing the DTM algorithm in an accessible form.

The Kenya Ministry of Health is acknowledged for maintaining the public health facility registry, and the Humanitarian Data Exchange (HDX) for curating and distributing this critical infrastructure dataset.

## Notes

### Competing Interest Statement

The authors have declared no competing interest.

### Summary of Updates

Title and version updated throughout. Abstract expanded to cover the three new additions and 91% temporal classification accuracy; keywords extended. Contributions grew from 5 to 8 items. Introduction gained one paragraph on operational motivation. Three new Methods subsections added: Observed-to-Expected Completeness Engine: expected case count derivation, four-class severity scheme, seven-disease WHO reference rate table (Table 2), deficit rate as DTM point-cloud weight. Causal Classification of Detected Voids: five-class decision tree (BORDER / ACCESS / INFRASTRUCTURE / SYSTEM / UNKNOWN) formalized as a piecewise equation with covariate triggers and operational interpretations for each class. Temporal Void Classification: Fano factor derivation, two-state HMM (Baum-Welch EM + Viterbi decoding), structural probability formula, three-class decision rule with Fano-based overrides, and Table 3 summarizing labels and recommended actions. Algorithm box extended with three new annotated blocks: O/E engine loop, causal labelling call, and temporal classification loop (conditional on T ≥ 2 periods). Two new Results subsections added: Temporal Classifier Validation: 91% overall accuracy, per-class recall (STRUCTURAL 0.94, STOCHASTIC 0.89, INTERMITTENT 0.85), Fano override impact (+3pp STRUCTURAL recall), HMM convergence characterization. Causal Classification Performance: 78% accuracy, κ = 0.71 (95% CI: 0.63 to 0.79), per-class precision/recall analysis, SYSTEM class identified as hardest without DHIS2 audit logs. Nyanza case study table split into two sub-tables to fit page margins. P(structural) values added for each void. Discussion updated throughout. Principal Findings reflects six advances rather than four. Limitations adds two bullet points (temporal classification requires ≥ 2 periods; causal labels are decision-tree approximations requiring field validation). Future Work adds DHIS2 real-time integration and extended causal covariates. Conclusion rewritten to reflect expanded scope. Data Availability reference updated from Tables 1 to 3 to Tables 1 to 5.

## References

[1] Silverman, B. W. (1986). Density Estimation for Statistics and Data Analysis. Chapman and Hall. DOI: 10.1007/978-1-4899-3324-9

[2] Okabe, A., Boots, B., Sugihara, K., & Chiu, S. N. (2009). Spatial Tessellations: Concepts and Applications of Voronoi Diagrams (2nd ed.). John Wiley & Sons. DOI: 10.1002/9780470317013

[3] Chazal, F., Cohen-Steiner, D., & Mérigot, Q. (2011). Geometric inference for probability measures. Foundations of Computational Mathematics, 11(6), 733–751. DOI: 10.1007/s10208-011-9098-0

[4] Kulldorff, M. (1997). A spatial scan statistic. Communications in Statistics—Theory and Methods, 26(6), 1481–1496. DOI: 10.1080/03610929708831995

[5] Kulldorff, M. (2023). SaTScan™: Software for the spatial, temporal, and space-time scan statistics. URL: https://www.satscan.org/

[6] Skaf, Y., & Laubenbacher, R. (2022). Topological data analysis in biomedicine: A review. Journal of Biomedical Informatics, 130, 104082. DOI: 10.1016/j.jbi.2022.104082

[7] Lum, P. Y., Singh, G., Lehman, A., Ishkanov, T., Vejdemo-Johansson, M., Alagappan, M., Carlsson, J., & Carlsson, G. (2013). Extracting insights from the shape of complex data using topology. Scientific Reports, 3, 1236. DOI: 10.1038/srep01236

[8] Edelsbrunner, H., & Harer, J. (2010). Computational Topology: An Introduction. American Mathematical Society. DOI: 10.1090/mbk/069

[9] Chen, Y.-C., Genovese, C. R., & Wasserman, L. (2014). Enhanced mode clustering. Journal of the American Statistical Association, 111(514), 704–719. DOI: 10.1080/01621459.2016.1165103 arXiv: 1406.1780

[10] Welzl, E. (1991). Smallest enclosing disks (balls and ellipsoids). In H. Maurer (Ed.), New Results and New Trends in Computer Science (pp. 359–370). Springer. DOI: 10.1007/BFb0038202

[11] Satopää, V., Albrecht, J., Irwin, D., & Raghavan, B. (2011). Finding a “kneedle” in a haystack: Detecting knee points in system behavior. In 2011 31st International Conference on Distributed Computing Systems Workshops (pp. 166–171). IEEE. DOI: 10.1109/ICDCSW.2011.20

[12] Jaccard, P. (1901). Étude comparative de la distribution florale dans une portion des Alpes et du Jura. Bulletin de la Société Vaudoise des Sciences Naturelles, 37, 547–579. DOI: 10.5169/seals-266450

[13] Davison, A. C., & Hinkley, D. V. (1997). Bootstrap Methods and their Application. Cambridge University Press. DOI: 10.1017/CBO9780511802843

